# Efficacy and Safety of Direct Oral Anticoagulants Versus Warfarin in Patients with Non-valvular Atrial Fibrillation and Liver Cirrhosis

**DOI:** 10.1101/2023.12.05.23299538

**Authors:** Tien-Shin Chou, Yuan Lin, Ming-Lung Tsai, Chin-Ju Tseng, Jhih-Wei Dai, Ning-I Yang, Chih-Lang Lin, Li-Wei Chen, Ming-Jui Hung, Tien-Hsing Chen

## Abstract

**Background:** Evidence of the pharmacodynamics of direct oral anticoagulants (DOACs) is limited in patients with atrial fibrillation (AF) and liver cirrhosis (LC). This study aimed to compare the efficacy and safety of DOACs versus warfarin in patients with non-valvular AF and LC.

**Methods:** We conducted a new-user, retrospective cohort study involving patients with AF and LC from the Chang Gung Hospital System for the years 2012–2021. LC was categorized per the Child–Pugh classification system. We divided the included patients into two cohorts, namely a DOAC cohort and a warfarin cohort. The measured outcomes were thromboembolic events (ischemic stroke [IS], transient ischemic attack [TIA], and systemic embolism [SE]), intracranial hemorrhage [ICH], gastrointestinal (GI) and major bleeding, and all-cause mortality.

**Results:** In total, 478 DOAC users and 247 warfarin users were included in the analysis. DOACs and warfarin exhibited comparable efficacy in preventing thromboembolic events, namely IS (adjusted hazard ratio [aHR], 1.05; 95% confidence interval [CI], 0.42–2.61), TIA (aHR, 1.36; 95% CI, 0.18–10.31]), and SE (aHR, 0.49; 95% CI, 0.14–1.70). DOAC use was associated with a similar risk of ICH (aHR, 0.65; 95% CI, 0.26–1.59) and GI bleeding (aHR, 0.64; 95% CI, 0.39–1.03), a decreased risk of major bleeding (aHR, 0.64; 95% CI, 0.42–0.99), and a reduction in all-cause mortality (aHR, 0.73; 95% CI, 0.54–0.99). Patients with Child– Pugh class A classification exhibited a significant reduction in major bleeding risk in DOAC users (aHR, 0.48; 95% CI, 0.33–0.70); however, this reduction was nonsignificant for patients with class B or C classification (aHR, 0.77; 95% CI, 0.54−1.08)

**Conclusion:** Relative to warfarin, DOACs provide comparable efficacy but greater safety for patients with non-valvular AF and LC. Specifically, DOAC use leads to a lower risk of major bleeding and a lower all-cause mortality.

**Clinical Perspective:** *What is New?:*

- This study reveals that direct oral anticoagulants (DOACs) and warfarin have similar effectiveness in preventing thromboembolic events in patients with non-valvular atrial fibrillation (AF) and liver cirrhosis (LC).
- DOACs demonstrate a lower risk of major bleeding and reduced all-cause mortality compared to warfarin, especially in patients with Child–Pugh class A LC.
- The safety profile of DOACs in reducing gastrointestinal bleeding is comparable to that of warfarin, with a trend towards lower risk.

*What are the Clinical Implications?:*

- For patients with non-valvular AF and LC, particularly those with Child–Pugh class A classification, DOACs may be a safer alternative to warfarin due to their lower risk of major bleeding.
- Clinicians can consider DOACs as a comparable alternative to warfarin for stroke prevention in this patient group, given their similar efficacy in preventing thromboembolic events.
- The reduced all-cause mortality associated with DOACs highlights their potential benefit in improving overall patient outcomes in the context of AF and LC.

## Introduction

Liver cirrhosis (LC) is a risk factor for atrial fibrillation (AF), and the prevalence rate of AF increases with the severity of liver disease.(1, 2). In AF management, anticoagulation therapy is a crucial component. However, when AF and LC coexist, a key challenge for implementing anticoagulation therapy is balancing the benefits of stroke prevention against the increased risk of bleeding; this challenge is related to the multifaceted hemostatic changes and altered drug metabolism induced by liver disease (3, 4).

In the general AF population, direct oral anticoagulants (DOACs) have been demonstrated to be effective for stroke prevention, and they pose a lower risk of bleeding compared with warfarin (5). However, the efficacy and safety of DOACs in patients with both AF and LC are less clear. DOACs provide several advantages, including simplified dosing adjustments based on renal function, limited drug–diet interactions, and minimal coagulation monitoring requirements (6). Because DOACs are less dependent on liver clearance relative to warfarin, they can be advantageous for patients with LC (7). However, evidence of the pharmacodynamics of DOACs in patients with LC is limited because most clinical trials exclude patients with severe liver disease (8–11).

On the basis of the Child–Pugh classification system, current guidelines advocate the administration of oral anticoagulants for patients with LC (12–14). For patients with class A LC, all four currently available DOACs are regarded as appropriate. For those with class B LC, dabigatran, apixaban, and edoxaban should be administered with caution, whereas the use of rivaroxaban is discouraged. For patients with class C LC, DOACs are not recommended (12–14). Nonetheless, most of the data supporting these guidelines were derived from small-scale clinical studies (15, 16) or extrapolated from noncirrhotic populations(17–20). That is, these data were not based on comprehensive clinical and laboratory information with accurate definitions of liver disease severity (17–20). Furthermore, real-world studies have yielded inconclusive and inconsistent data regarding the effectiveness and safety of DOACs in patients with liver disease (17–19). Lee SR et al. reported that the clinical outcomes of DOAC and warfarin use in patients with LC were not significantly different (17). By contrast, Lee HF et al. reported a lower risk of gastrointestinal (GI) bleeding in DOAC users than in warfarin users (18), and Lawal et al. observed that relative to warfarin users, DOAC users were exposed to a comparable risk of major GI bleeding but to lower risks of all-cause mortality and major bleeding (19). These inconsistent findings and the limited event data pertaining to this topic highlight the necessity of conducting more focused and rigorous research in this area.

Thus, we enrolled cohorts of patients with both AF and LC who were treated with either DOACs or warfarin. Our objective was to investigate the comparative efficacy and safety of DOACs versus warfarin in patients with AF across the spectrum of LC severity.

## Methods

### Data source

The data were retrieved from the Chang Gung Research Database (21). Due to retrospective nature of the database study, the hospital identification number of each patient was encrypted and de-identified to protect their privacy. Therefore, informed consent was waived for this study. The institutional review board of Chang Gung Memorial Hospital approved the study protocol (IRB No. :202200625B0).

### Study Population and Exposure

The study cohort comprised patients with both nonvalvular AF and LC who were exposed to DOACs (apixaban, dabigatran, rivaroxaban, or edoxaban) or warfarin between January 1, 2012, and December 31, 2021. The index day was defined as the first day on which a DOAC or warfarin was prescribed. Patients with LC were included if they met two criteria. First, they were required to be clinically diagnosed with LC. Second, this diagnosis was required to be validated through a manual review of reports of abdominal ultrasound or transient elastography (FibroScan, liver stiffness > 12 kPa) conducted 3 years before or 6 months after the index day (22, 23). Patients were excluded if they lacked abdominal ultrasound or transient elastography data, their abdominal ultrasound or transient elastography results did not provide any evidence of LC, they had a history of liver transplantation, their demographic (age and sex) or Child–Pugh score data were incomplete, they were aged <20 years, they had valvular AF, they had prior venous thromboembolism, they were undergoing dialysis for end-stage renal disease, or they were lost to follow-up. Furthermore, we divided the included patients into a DOAC group and a warfarin group (**Figure 1**).

**Figure 1.**
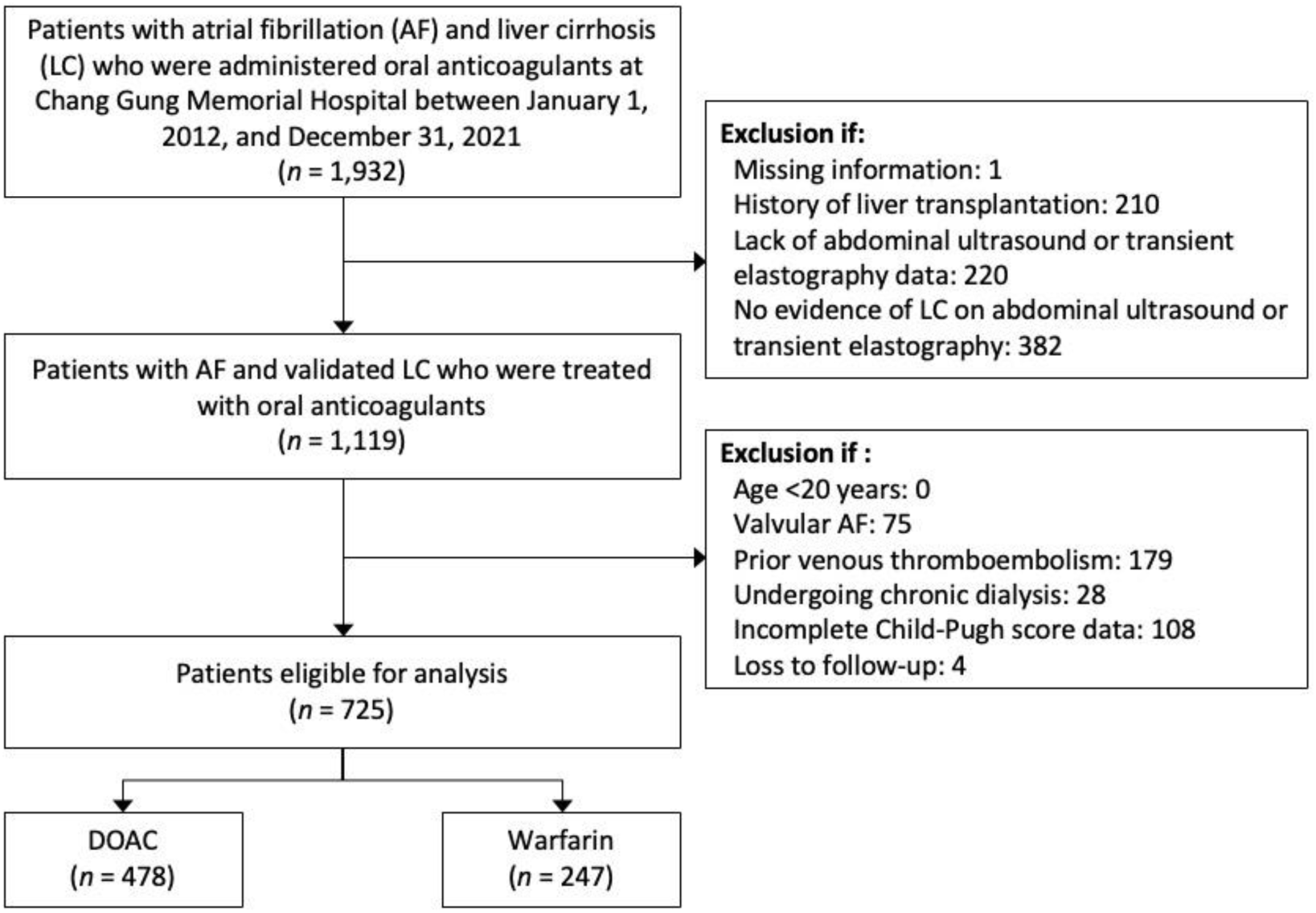
Flow chart for selecting and excluding patients.

### Clinical outcomes

The primary outcomes were efficacy and safety outcomes. The efficacy outcomes were ischemic stroke (IS), transient ischemic attack (TIA), systemic embolism (SE), and a composite measure of IS, TIA, and SE (IS/TIA/SE). The safety outcomes were intracranial hemorrhage (ICH), GI bleeding, and major bleeding. The secondary outcomes were all-cause mortality, myocardial infarction (MI), and liver decompensation outcomes. The liver decompensation outcomes were hepatic encephalopathy, ascites, spontaneous bacterial infection, hepatorenal syndrome, esophageal variceal bleeding, hepatocellular carcinoma (HCC), and liver related death. Comorbidities and diseases were detected by referencing International Classification of Diseases, 9th and 10th Revision, Clinical Modification (ICD-9-CM and ICD-10-CM) codes **(Supplemental Table S1)**. Most of the clinical events were identified through inpatient diagnoses, with the exceptions being ascites, and all-cause mortality. Information on the date and cause of death was retrieved from the Taiwan Death Registry database. Ascites was defined as newly detectable ascites that were clinically confirmed by ultrasound findings. The end of the study period was the date of death, December 31, 2021, the final clinical visit, or the fifth year of follow-up, whichever occurred first.

### Covariate measurement

The covariates examined in the present study were demographics (i.e., age, sex, smoking habit, alcohol drinking habit, and body mass index), duration of LC, LC-related diseases (e.g., primary biliary cholangitis), LC complications (e.g., ascites), comorbidities (i.e., diabetes mellitus, hypertension, and four others), historical events requiring hospitalization (i.e., ischemic stroke, major bleeding, and six others), medications concurrently prescribed at outpatient clinics (i.e., aspirin, beta-blockers, and 16 other medications), risk scores (i.e., Child–Pugh score, CHA2DS2-VASc score, HAS-BLED score, and Charlson comorbidity index score), liver function test (i.e., serum aspartate aminotransferase and alanine aminotransferase, platelet count, prothrombin time, bilirubin level, and albumin level), and various laboratory data (e.g., international normalized ratio [INR]). A comorbidity was included in this study if two outpatient diagnoses or one discharge diagnosis was made for the comorbidity prior to the index date. Laboratory data within 3 months prior to the index date were retrieved. Liver function test results and esophagogastroduodenoscopy results were retrieved from records dating to 3 years prior to and 6 months after the index date. Concurrent medication use was determined using records dating to 3 months before or after the index date. Portal hypertension was defined as the presence of ascites, varices, or splenomegaly (24). The classification of liver disease severity based on Child–Pugh scores is described in **Supplemental Table S2**.

### Statistical analysis

Because substantial differences in baseline demographics and clinical characteristics were identified between the DOAC and warfarin groups, inverse probability of treatment weighting (IPTW), which is based with on the average treatment effect and propensity scores, was performed to ensure comparability between the two groups. We estimated the propensity score using a generalized boosted model (GBM), which was constructed using 50,000 regression trees. All the covariates (**Table 1**) were included in the estimation of propensity scores; the follow-up year was replaced by the index date. The balance of baseline data between the two groups before and after GBM-IPTW was evaluated by calculating standardized difference (STD), where an absolute STD value of <0.2 indicated a non-substantial group difference. Because of the substantial amount of missing values in the continuous laboratory data, the missing data were imputed using an expectation-maximization algorithm. We compared outcomes in both the imputed and IPTW-adjusted cohorts.

**Table 1.** Baseline characteristics of patients with atrial fibrillation and liver cirrhosis who were administered DOAC or warfarin.

| Variable | Before IPTW and imputation# | | | | After IPTW and imputation\$ | | |
| --- | --- | --- | --- | --- | --- | --- | --- |
|  | Available number | DOAC (n = 478) | Warfarin (n = 247) | STD | DOAC | Warfarin | STD |
| Age, years | 725 | 73.2 ± 10.9 | 65.2 ± 12.6 | 0.68 | 72.2 ± 11.3 | 67.8 ± 12.4 | 0.37 |
| Age group | 725 |  |  |  |  |  |  |
| <65 years |  | 103 (21.5) | 122 (49.4) | -0.61 | 24.8% | 39.3% | -0.31 |
| 65–74 years |  | 147 (30.8) | 64 (25.9) | 0.11 | 30.6% | 30.2% | 0.01 |
| ≥75 years |  | 228 (47.7) | 61 (24.7) | 0.49 | 44.6% | 30.5% | 0.29 |
| Female | 725 | 181 (37.9) | 82 (33.2) | 0.10 | 37.6% | 33.0% | 0.10 |
| Smoking | 725 | 133 (27.8) | 80 (32.4) | -0.10 | 29.0% | 29.1% | <0.01 |
| Alcohol drinking | 725 | 112 (23.4) | 63 (25.5) | -0.05 | 25.1% | 23.8% | 0.03 |
| Body mass index, kg/m<sup>2</sup> | 615 | 26.1 ± 4.9 | 25.5 ± 4.9 | 0.12 | 26.2 ± 4.6 | 25.6 ± 4.5 | 0.12 |
| Vital signs |  |  |  |  |  |  |  |
| Systolic blood pressure, mmHg | 669 | 131.2 ± 21.9 | 130.7 ± 22.3 | 0.03 | 131.0 ± 21.2 | 132.2 ± 22.9 | -0.05 |
| Diastolic blood pressure, mmHg | 669 | 75.2 ± 13.2 | 73.3 ± 13.4 | 0.14 | 75.2 ± 12.7 | 73.7 ± 13.4 | 0.11 |
| Heart rate, beats/min | 667 | 83.5 ± 18.0 | 81.2 ± 18.2 | 0.13 | 83.5 ± 17.2 | 80.3 ± 17.2 | 0.19 |
| Duration of liver cirrhosis, years | 725 | 4.3 ± 5.2 | 3.5 ± 4.1 | 0.19 | 4.1 ± 5.0 | 3.8 ± 4.6 | 0.07 |
| Liver cirrhosis related disease |  |  |  |  |  |  |  |
| Alcoholism | 725 | 79 (16.5) | 39 (15.8) | 0.02 | 17.6% | 13.2% | 0.12 |
| HBV infection | 725 | 153 (32.0) | 75 (30.4) | 0.04 | 32.0% | 33.6% | -0.04 |
| HCV infection | 725 | 152 (31.8) | 78 (31.6) | <0.01 | 30.4% | 31.1% | -0.02 |
| Nonalcoholic fatty liver disease | 725 | 46 (9.6) | 16 (6.5) | 0.12 | 9.3% | 6.5% | 0.10 |
| Autoimmune hepatitis | 725 | 5 (1.0) | 0 (0.0) | 0.15 | 1.0% | 0.0% | 0.14 |
| Primary biliary cholangitis | 725 | 6 (1.3) | 1 (0.4) | 0.09 | 1.3% | 0.5% | 0.08 |
| Liver cirrhosis complications |  |  |  |  |  |  |  |
| Ascites | 725 | 147 (30.8) | 99 (40.1) | -0.20 | 32.5% | 36.8% | -0.09 |
| EV or GV | 725 | 62 (13.0) | 56 (22.7) | -0.26 | 13.7% | 21.9% | -0.22 |
| Hepatic encephalopathy | 725 | 10 (2.1) | 15 (6.1) | -0.20 | 2.3% | 5.0% | -0.14 |
| Spontaneous bacterial peritonitis | 725 | 2 (0.4) | 1 (0.4) | <0.01 | 0.4% | 1.1% | -0.08 |
|  | Available number | DOAC (n = 478) | Warfarin (n = 247) | STD | DOAC | Warfarin | STD |
| HCC | 725 | 123 (25.7) | 56 (22.7) | 0.07 | 24.3% | 23.6% | 0.02 |
| Hepatorenal syndrome | 725 | 4 (0.8) | 1 (0.4) | 0.06 | 0.8% | 0.4% | 0.06 |
| Portal hypertension | 725 | 350 (73.2) | 216 (87.4) | -0.36 | 74.3% | 85.2% | -0.27 |
| Treatment for HCC |  |  |  |  |  |  |  |
| Systemic therapy | 725 | 66 (13.8) | 37 (15.0) | -0.03 | 13.0% | 16.3% | -0.09 |
| Liver resection | 725 | 5 (1.0) | 6 (2.4) | -0.11 | 0.9% | 2.8% | -0.14 |
| Radiofrequency ablation | 725 | 75 (15.7) | 21 (8.5) | 0.22 | 15.1% | 8.2% | 0.21 |
| Transarterial chemoembolisation | 725 | 8 (1.7) | 0 (0.0) | 0.18 | 1.5% | 0.0% | 0.17 |
| Percutaneous injection therapy | 725 | 11 (2.3) | 8 (3.2) | -0.06 | 2.1% | 3.1% | -0.06 |
| Comorbidities |  |  |  |  |  |  |  |
| Diabetes mellitus | 725 | 219 (45.8) | 110 (44.5) | 0.03 | 46.3% | 48.0% | -0.03 |
| Hypertension | 725 | 368 (77.0) | 152 (61.5) | 0.34 | 75.9% | 68.1% | 0.18 |
| Hyperlipidemia | 725 | 147 (30.8) | 61 (24.7) | 0.14 | 30.2% | 26.1% | 0.09 |
| Peripheral artery disease | 725 | 45 (9.4) | 34 (13.8) | -0.14 | 9.1% | 13.0% | -0.12 |
| Peptic ulcer disease | 725 | 242 (50.6) | 123 (49.8) | 0.02 | 50.7% | 51.4% | -0.01 |
| Malignancy | 725 | 222 (46.4) | 101 (40.9) | 0.11 | 45.0% | 43.4% | 0.03 |
| History of events requiring hospitalization |  |  |  |  |  |  |  |
| Systemic thromboembolism | 725 | 11 (2.3) | 13 (5.3) | -0.16 | 2.3% | 4.5% | -0.12 |
| Ischemic stroke | 725 | 71 (14.9) | 31 (12.6) | 0.07 | 15.6% | 10.1% | 0.17 |
| Transient ischemic attack | 725 | 8 (1.7) | 10 (4.0) | -0.14 | 1.5% | 2.7% | -0.09 |
| Intracranial hemorrhage | 725 | 31 (6.5) | 7 (2.8) | 0.17 | 5.8% | 2.4% | 0.18 |
| Myocardial infarction | 725 | 26 (5.4) | 9 (3.6) | 0.09 | 5.3% | 3.5% | 0.09 |
| Coronary revascularization | 725 | 32 (6.7) | 11 (4.5) | 0.10 | 6.4% | 3.2% | 0.15 |
| Heart failure | 725 | 122 (25.5) | 71 (28.7) | -0.07 | 26.1% | 27.9% | -0.04 |
| Major bleeding | 725 | 122 (25.5) | 48 (19.4) | 0.15 | 24.6% | 20.9% | 0.09 |
| Medications |  |  |  |  |  |  |  |
| Aspirin | 725 | 126 (26.4) | 54 (21.9) | 0.11 | 27.4% | 21.0% | 0.15 |
|  | Available number | DOAC (n = 478) | Warfarin (n = 247) | STD | DOAC | Warfarin | STD |
| P2Y12 | 725 | 75 (15.7) | 17 (6.9) | 0.28 | 15.6% | 7.6% | 0.25 |
| Amiodarone/dronedarone | 725 | 72 (15.1) | 30 (12.1) | 0.09 | 14.9% | 12.7% | 0.06 |
| Digoxin | 725 | 47 (9.8) | 51 (20.6) | -0.30 | 10.3% | 17.1% | -0.20 |
| ACEi/ARBs | 725 | 244 (51.0) | 104 (42.1) | 0.18 | 50.9% | 44.1% | 0.14 |
| Beta-blockers |  |  |  |  |  |  |  |
| Propranolol | 725 | 33 (6.9) | 29 (11.7) | -0.17 | 7.0% | 11.2% | -0.15 |
| Carvedilol | 725 | 29 (6.1) | 21 (8.5) | -0.09 | 6.2% | 6.6% | -0.02 |
| Other beta-blockers | 725 | 191 (40.0) | 50 (20.2) | 0.44 | 38.2% | 23.5% | 0.32 |
| Calcium channel blockers | 725 | 175 (36.6) | 91 (36.8) | <0.01 | 36.0% | 35.3% | 0.01 |
| MRAs | 725 | 126 (26.4) | 88 (35.6) | -0.20 | 27.6% | 35.1% | -0.16 |
| Loop diuretics | 725 | 235 (49.2) | 129 (52.2) | -0.06 | 49.7% | 51.2% | -0.03 |
| Thiazide | 725 | 17 (3.6) | 14 (5.7) | -0.10 | 3.8% | 4.4% | -0.03 |
| Antiviral treatment for HBV or HCV | 725 | 91 (19.0) | 33 (13.4) | 0.15 | 19.1% | 12.7% | 0.18 |
| PPIs | 725 | 148 (31.0) | 86 (34.8) | -0.08 | 32.1% | 35.2% | -0.07 |
| Ursodeoxycholic acid | 725 | 14 (2.9) | 4 (1.6) | 0.09 | 2.7% | 1.3% | 0.10 |
| NSAIDs | 725 | 129 (27.0) | 71 (28.7) | -0.04 | 27.1% | 25.4% | 0.04 |
| Hypoglycemic agent |  |  |  |  |  |  |  |
| SGLT2i | 725 | 11 (2.3) | 1 (0.4) | 0.16 | 2.1% | 0.4% | 0.15 |
| GLP1RA | 725 | 3 (0.6) | 0 (0.0) | 0.11 | 0.5% | 0.0% | 0.10 |
| Other oral hypoglycemic agents | 725 | 144 (30.1) | 61 (24.7) | 0.12 | 31.0% | 26.7% | 0.10 |
| Insulin | 725 | 58 (12.1) | 35 (14.2) | -0.06 | 13.1% | 14.3% | -0.04 |
| Risk score |  |  |  |  |  |  |  |
| CHA<sub>2</sub>DS<sub>2</sub>-VASc | 725 | 3.6 ± 1.7 | 2.9 ± 1.9 | 0.39 | 3.5 ± 1.7 | 3.1 ± 1.8 | 0.26 |
| HAS-BLED | 725 | 3.9 ± 1.3 | 3.6 ± 1.5 | 0.17 | 3.9 ± 1.3 | 3.6 ± 1.4 | 0.20 |
| Charlson comorbidity index | 725 | 6.0 ± 3.3 | 5.6 ± 3.2 | 0.10 | 5.9 ± 3.3 | 5.8 ± 3.1 | 0.01 |
| Child–Pugh score | 725 |  |  |  |  |  |  |
| Class A: 5 or 6 |  | 265 (55.4) | 94 (38.1) | 0.35 | 53.9% | 45.3% | 0.17 |
|  | Available number | DOAC (n = 478) | Warfarin (n = 247) | STD | DOAC | Warfarin | STD |
| Class B: 7–9 |  | 190 (39.7) | 135 (54.7) | -0.30 | 40.5% | 47.4% | -0.14 |
| Class C: ≥10 |  | 23 (4.8) | 18 (7.3) | -0.10 | 5.6% | 7.3% | -0.07 |
| Baseline laboratory data |  |  |  |  |  |  |  |
| eGFR, mL/min/1.73 m<sup>2</sup> | 709 | 73.3 ± 33.3 | 79.4 ± 45.4 | -0.15 | 74.0 ± 33.7 | 78.9 ± 41.7 | -0.13 |
| eGFR status | 709 |  |  |  |  |  |  |
| >60 |  | 306 (64.6) | 155 (66.0) | -0.03 | 64.7% | 67.6% | -0.06 |
| 30–60 |  | 139 (29.3) | 54 (23.0) | 0.14 | 29.2% | 23.7% | 0.13 |
| <30 |  | 29 (6.1) | 26 (11.1) | -0.18 | 6.1% | 8.7% | -0.10 |
| Serum creatinine, µmol/L | 709 | 106.1 ± 79.6 | 132.6 ± 150.3 | -0.19 | 106.1 ± 79.6 | 123.8 ± 123.8 | -0.13 |
| Blood urea nitrogen, mmol/L | 610 | 8.2 ± 5.5 | 9.2 ± 7.8 | -0.15 | 7.9 ± 5.3 | 8.7 ± 6.5 | -0.13 |
| AST, U/L | 668 | 43.8 ± 36.7 | 50.5 ± 38.2 | -0.18 | 44.0 ± 37.7 | 46.4 ± 33.8 | -0.07 |
| ALT, U/L | 715 | 31.5 ± 27.5 | 39.9 ± 46.9 | -0.22 | 32.5 ± 29.0 | 37.0 ± 38.8 | -0.13 |
| ALK-P, U/L | 484 | 104.2 ± 78.6 | 106.1 ± 58.3 | -0.03 | 99.4 ± 68.1 | 107.4 ± 58.3 | -0.13 |
| Albumin level, g/L | 725 | 35.9 ± 6.6 | 35.8 ± 6.3 | 0.02 | 35.7 ± 6.6 | 35.5 ± 6.1 | 0.03 |
| Total bilirubin level, µmol/L | 725 | 20.4 ± 17.8 | 23.2 ± 36.0 | -0.10 | 21.3 ± 21.3 | 22.5 ± 30.6 | -0.04 |
| Total bilirubin status, µmol/L | 725 |  |  |  |  |  |  |
| <34.2 |  | 432 (90.4) | 217 (87.9) | 0.08 | 89.5% | 87.0% | 0.08 |
| ≥34.2 |  | 46 (9.6) | 30 (12.1) | -0.08 | 10.5% | 13.0% | -0.08 |
| International normalized ratio (INR) | 725 | 1.2 ± 0.3 | 1.5 ± 0.6 | -0.63 | 1.2 ± 0.3 | 1.4 ± 0.5 | -0.35 |
| INR status | 725 |  |  |  |  |  |  |
| <1.7 |  | 455 (95.2) | 180 (72.9) | 0.64 | 93.6% | 80.8% | 0.39 |
| 1.7–2.3 |  | 20 (4.2) | 36 (14.6) | -0.36 | 5.3% | 11.6% | -0.23 |
| ≥2.3 |  | 3 (0.6) | 31 (12.6) | -0.50 | 1.1% | 7.6% | -0.33 |
| Hemoglobin, g/L | 678 | 120.0 ± 23.8 | 117.6 ± 24.1 | 0.10 | 120.3 ± 23.6 | 119.5 ± 23.6 | 0.03 |
| Platelet, count ×10<sup>3</sup> | 669 | 161.8 ± 79.7 | 157.0 ± 114.7 | 0.05 | 162.6 ± 77.2 | 161.3 ± 103.0 | 0.01 |
| Fasting glucose, mmol/L | 312 | 6.6 ± 2.1 | 6.9 ± 3.1 | -0.13 | 6.6 ± 2.0 | 6.6 ± 2.9 | <0.01 |
| Glycated hemoglobin, % | 426 | 6.4 ± 1.3 | 6.7 ± 1.4 | -0.23 | 6.2 ± 1.2 | 6.3 ± 1.3 | -0.07 |

| Variable | Before IPTW and imputation<sup>#</sup> | | | | After IPTW and imputation<sup>\$</sup> | | |
| --- | --- | --- | --- | --- | --- | --- | --- |
|  | Available number | DOAC (n = 478) | Warfarin (n = 247) | STD | DOAC | Warfarin | STD |
| Triglyceride, mmol/L | 449 | 1.1 ± 0.7 | 1.2 ± 0.8 | -0.15 | 1.1 ± 0.6 | 1.1 ± 0.6 | <0.01 |
| Total cholesterol, mmol/L | 474 | 3.9 ± 1.0 | 4.1 ± 1.3 | -0.16 | 3.9 ± 1.0 | 4.1 ± 1.1 | -0.14 |
| High-density lipoprotein, mmol/L | 420 | 1.1 ± 0.4 | 1.1 ± 0.5 | -0.05 | 1.1 ± 0.4 | 1.2 ± 0.4 | -0.15 |
| Low-density lipoprotein, mmol/L | 454 | 2.3 ± 0.9 | 2.4 ± 1.1 | -0.08 | 2.3 ± 0.8 | 2.4 ± 0.9 | -0.11 |
| Follow-up years | 725 | 2.4 ± 2.0 | 3.6 ± 2.9 | -0.48 | 2.6 ± 2.2 | 3.2 ± 2.6 | -0.25 |

Abbreviation: GBM, generalized boosted model; IPTW, inverse probability of treatment weighting; DOAC, direct oral anticoagulant; STD, standardized difference; EGD, esophagogastroduodenoscopy; EV, esophageal varices; GV, gastric varices; HBV, hepatitis B virus; HCV, hepatitis C virus; HCC, hepatocellular carcinoma; ACEi, angiotensin converting enzyme inhibitor; ARB, angiotensin receptor blockers; MRAs, mineralocorticoid receptor antagonists; PPIs, proton pump inhibitors; NSAIDs, nonsteroidal anti-inflammatory drugs; SGLT2i, sodium–glucose cotransporter 2 inhibitors; GLP1RA, glucagon-like peptide 1 receptor agonists; eGFR, estimated glomerular filtration rate; AST, aspartate aminotransferase; ALT, alanine aminotransferase; ALK-P, alkaline phosphatase.
<sup>#</sup> Data are presented as frequencies (percentages) and means ± standard deviations or medians [25th, 75th percentiles].
<sup>\$</sup> Data are presented as numbers (percentages) and means ± standard deviations or medians [25th, 75th percentiles].

We employed the Cox proportional hazard model to compare the risk of fatal time-to-event outcomes (e.g., liver-related death) between the DOAC and warfarin groups. The Fine–Gray sub-distribution hazard model was used to compare the incidence of nonfatal time-to-event outcomes (e.g., ischemic stroke and major bleeding) between the two groups; all-cause mortality during follow-up was considered as a competing risk in this model. Given the potential covariate imbalances between the two groups, even after thorough GBM-IPTW, including imbalances pertaining to age, age groups, esophageal/gastric varice–related complications, portal hypertension, radiofrequency ablation, P2Y12, other beta-blockers, CHA_2_DS_2_-VASc score, INR value, and INR status (absolute STD > 0.2), the aforementioned survival regression models were further adjusted for these covariates. Finally, a subgroup analysis of the primary safety outcome (i.e., major bleeding) was performed, and the results were stratified by clinical variables in the IPTW-adjusted cohort. SAS (version 9.4, SAS Institute) was used for statistical analyses. A two-sided *P* value of <0.05 was regarded as statistically significant.

## Results

### Baseline characteristics

From the Chang Gung Research Database, we identified 1,932 patients with AF and LC who were treated with oral anticoagulants between January 1, 2012, and December 31, 2021. Among them, 725 patients met the eligibility criteria for inclusion in the present study (**Figure 1**). **Table 1** presents the baseline characteristics of the study population, and the patients treated with DOACs (n = 478) were compared with those treated with warfarin (n = 247). After IPTW adjustment, the distribution of Child–Pugh classes was comparable between the DOAC group and the warfarin group (Child–Pugh A, 53.9% vs. 45.3%; STD = −0.17; Child–Pugh B, 40.5% vs. 47.4%; STD = −0.14; Child–Pugh C, 5.6% vs. 7.3%; STD = −0.07). However, the DOAC group had higher CHA2DS2-VASc scores (DOAC vs. warfarin, 3.5 vs. 3.1) and HAS-BLED scores (DOAC vs. warfarin, 3.9 vs. 3.6) relative to the warfarin group. Additionally, the DOAC group had a lower prevalence of portal hypertension (DOAC vs. warfarin, 74.3% vs. 85.2%), higher mean age (DOAC vs. warfarin, 72.2 years vs. 67.8 years), and lower INR level (DOAC vs. warfarin, 1.2 vs. 1.4) relative to the warfarin group. The average follow-up durations for the DOAC and warfarin groups were 2.6 and 3.2 years, respectively.

### Clinical outcomes

For the maximum 5-year follow-up period, the results revealed that DOACs were as effective as warfarin in reducing the incidence of the composite outcome (IS/TIA/SE; adjusted hazard ratio [aHR], 0.76; 95% confidence interval [CI], 0.38– 1.53) (**Table 2** and **Figure 2A**). The primary efficacy outcomes did not differ significantly between the two groups. Regarding safety outcomes, while there was no significant difference in intracranial hemorrhage (ICH) between the two groups, DOACs demonstrated a non-significant trend toward a lower risk of GI bleeding (aHR, 0.64; 95% CI, 0.39–1.03) and a significantly lower risk of major bleeding (aHR, 0.64; 95% CI, 0.42–0.99) relative to the warfarin group (Figure 2B). Furthermore, the DOAC group exhibited a significantly lower risk of all-cause mortality (aHR, 0.73; 95% CI, 0.54–0.99) (**Figure 2C**). The DOAC and warfarin groups exhibited similar levels of MI risk (aHR, 1.20; 95% CI, 0.26–5.61; *P* = 0.815). Regarding liver decompensation outcomes, no significant differences were observed between the two groups (**Table 2**). **Supplemental Figure S1** provides details on the causes of death.

**Figure 2.**
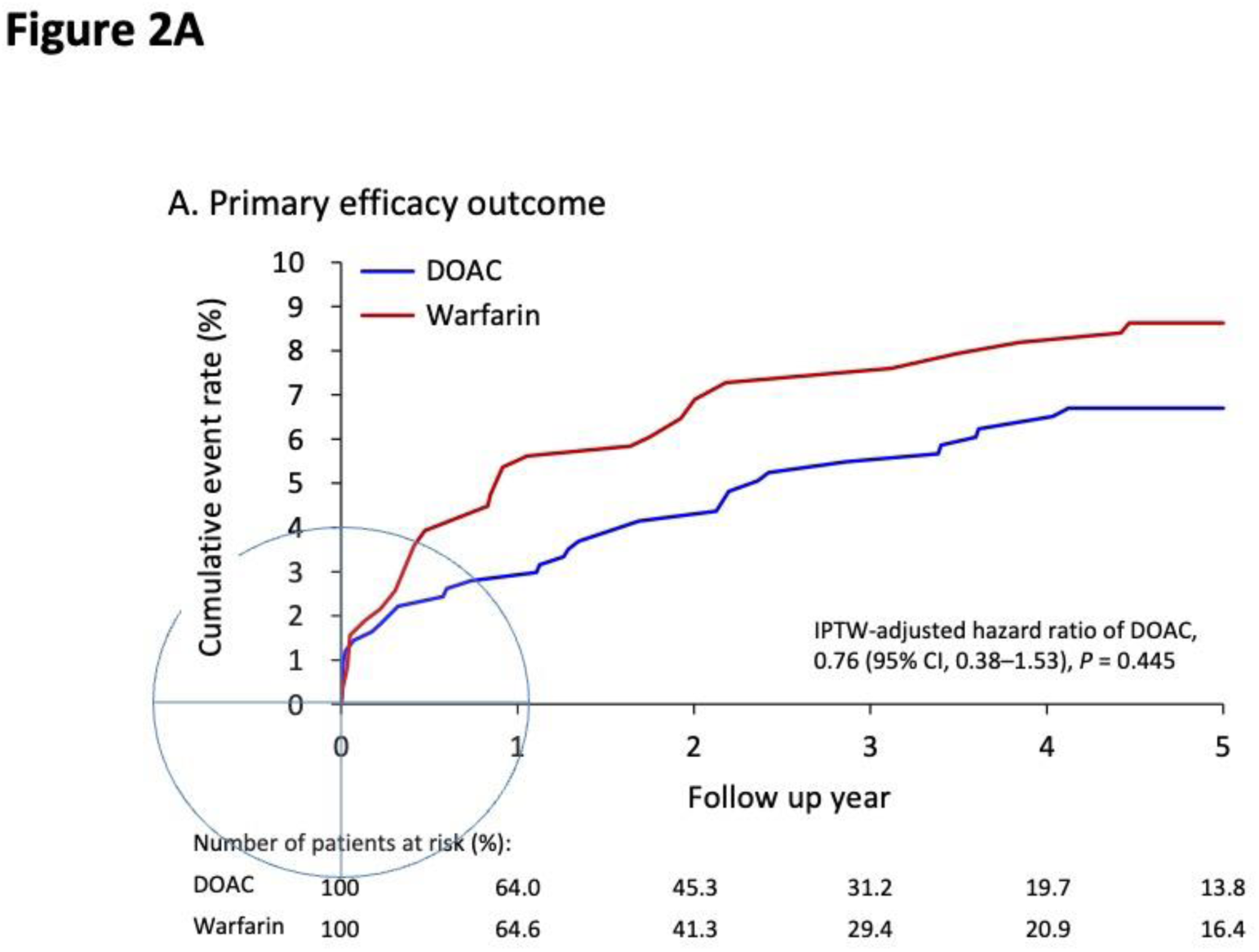
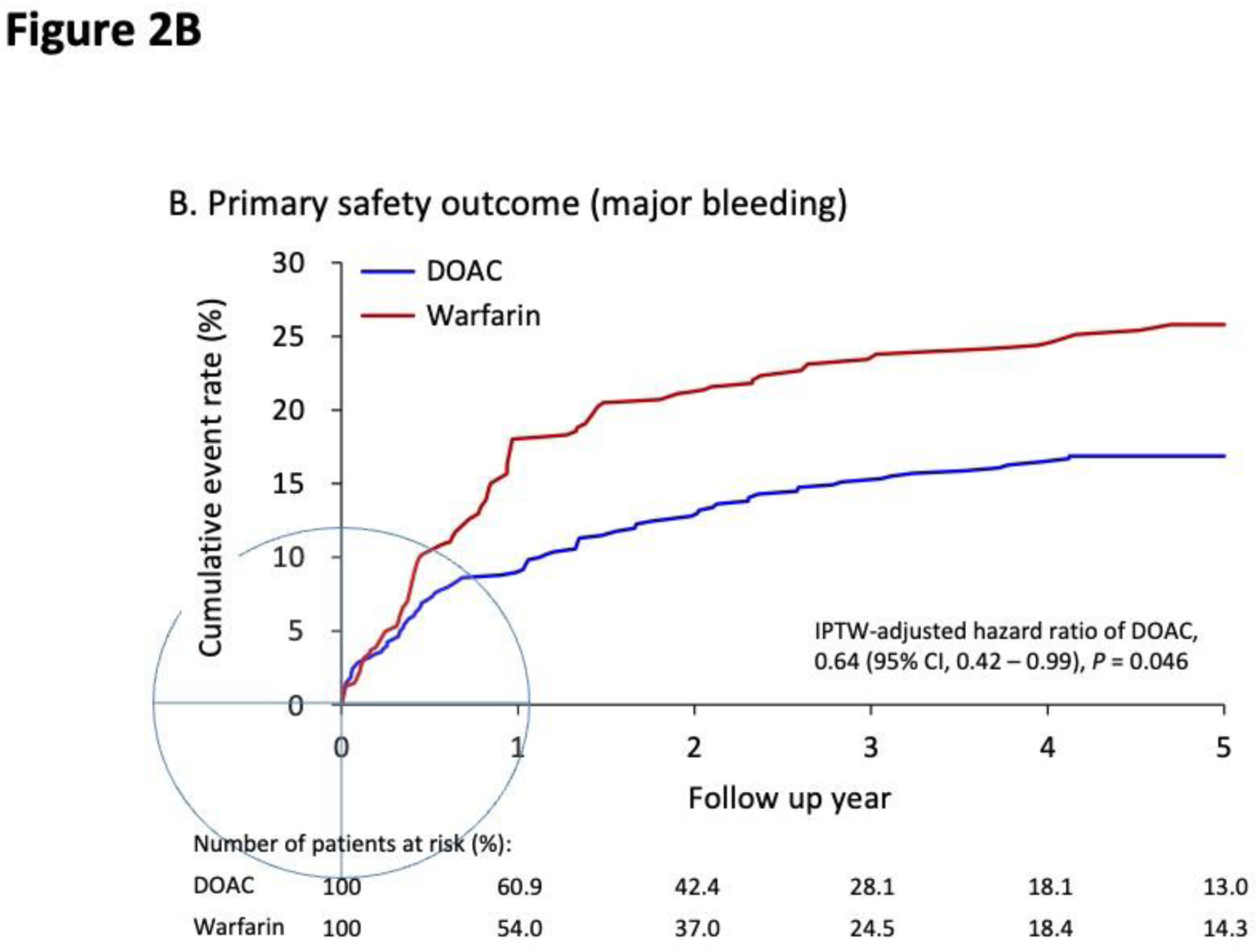
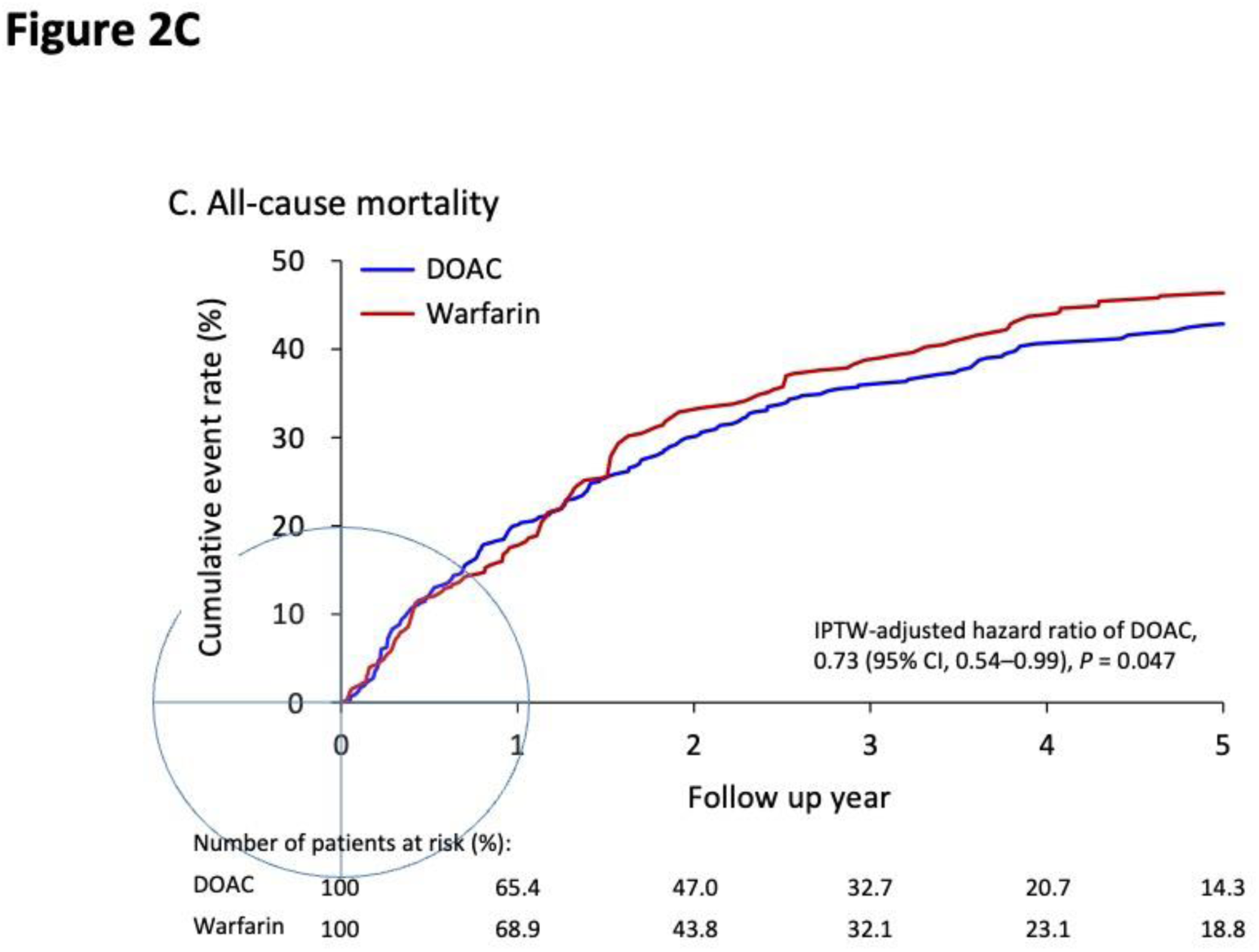
Cumulative event rates for primary efficacy outcome (A), major bleeding (B), and all-cause mortality (C) during 5-year follow-up period for patients with atrial fibrillation and liver cirrhosis who were administered DOACs or warfarin in IPTW-adjusted cohort. Abbreviations: DOAC, direct oral anticoagulant; IPTW, inverse probability of treatment weighting.

**Table 2.** Follow-up outcomes during 5-year follow-up of patients with atrial fibrillation and liver cirrhosis who were administered DOACs or warfarin in IPTW-adjusted cohort.

| Outcome | DOAC |  | Warfarin |  | aHR (95% CI)<br>for DOAC† | *P*<br>value |
| --- | --- | --- | --- | --- | --- | --- |
|  | Event rate | Incidence<br>(95% CI)* | Event rate | Incidence<br>(95% CI)* |  |  |
| Primary efficacy outcome |  |  |  |  |  |  |
| Ischemic stroke | 4.3% | 2.0 (1.2–2.7) | 4.7% | 2.2 (1.3–3.1) | 1.05 (0.42–2.61) | 0.921 |
| Transient ischemic attack | 0.7% | 0.3 (0.0–0.6) | 0.4% | 0.2 (–0.1–0.4) | 1.36 (0.18–10.3) | 0.764 |
| Systemic embolism | 1.9% | 0.8 (0.4–1.3) | 3.9% | 1.8 (1.0–2.6) | 0.49 (0.14–1.70) | 0.258 |
| Composite outcome | 6.7% | 3.1 (2.2–4.1) | 8.6% | 4.1 (2.9–5.4) | 0.76 (0.38–1.53) | 0.445 |
| Primary safety outcome |  |  |  |  |  |  |
| Intracranial hemorrhage | 3.3% | 1.5 (0.8–2.1) | 4.3% | 2.0 (1.1–2.8) | 0.65 (0.26–1.59) | 0.339 |
| Gastrointestinal bleeding | 13.1% | 6.4 (5.0–7.8) | 21.4% | 10.8 (8.8–12.9) | 0.64 (0.39–1.03) | 0.067 |
| Major bleeding | 16.9% | 8.4 (6.8–10.0) | 25.8% | 13.7 (11.3–16.1) | 0.64 (0.42–0.99) | 0.046 |
| Secondary outcomes |  |  |  |  |  |  |
| All-cause mortality | 42.9% | 19.4 (17.0–21.7) | 46.3% | 20.8 (18.1–23.5) | 0.73 (0.54–0.99) | 0.047 |
| Myocardial infarction | 1.9% | 0.9 (0.4–1.4) | 1.5% | 0.7 (0.2–1.1) | 1.20 (0.26–5.61) | 0.815 |
| Liver decompensation |  |  |  |  |  |  |
| Hepatic encephalopathy | 2.5% | 1.2 (0.6–1.8) | 4.9% | 2.2 (1.3–3.1) | NA | NA |
| New-onset ascites | 8.3% | 3.8 (2.8–4.9) | 10.9% | 5.1 (3.7–6.5) | 0.65 (0.34–1.23) | 0.183 |
| SBP | 2.2% | 1.0 (0.5–1.5) | 4.0% | 1.8 (1.0–2.6) | 0.69 (0.28–1.74) | 0.437 |
| Hepatorenal syndrome | 1.6% | 0.7 (0.3–1.2) | 4.6% | 2.1 (1.2–2.9) | NA | NA |
| Esophageal variceal bleeding | 3.4% | 1.6 (0.9–2.3) | 7.3% | 3.4 (2.3–4.6) | 0.65 (0.27–1.57) | 0.341 |
| New-onset HCC | 8.5% | 4.1 (3.0–5.2) | 6.2% | 3.0 (1.9–4.0) | 1.70 (0.77–3.78) | 0.192 |
| Liver-related death | 15.2% | 6.9 (5.5–8.3) | 21.3% | 9.6 (7.7–11.4) | 0.66 (0.38–1.15) | 0.143 |

Abbreviations: IPTW, inverse probability of treatment weighting; DOAC, direct oral anticoagulant; HR, hazard ratio; CI, confidence interval; NA, not acceptable; SBP, spontaneous bacterial infection; HCC, hepatocellular carcinoma.
† Additional adjustments were made for age, age group, esophageal/gastric varices–related complications, portal hypertension, radiofrequency ablation, P2Y12, other beta-blockers, CHA<sub>2</sub>DS<sub>2</sub>-VASc score, international normalized ratio, and INR status.
\* Number of events per 100 person-years.

### Subgroup analysis

In the subgroup analysis of major bleeding, the beneficial effect of DOACs in reducing major bleeding risk did not significantly differ across various factors, namely age, gender, portal hypertension, diabetes mellitus, malignancy, HAS-BLED score, albumin level, total bilirubin level, and INR status (**Supplemental Table S3**). However, the beneficial effect of DOACs was more in patients with HCC than in those without HCC (aHR, 0.38 vs. 0.72; *P* for interaction = 0.02). We further stratified our results by Child–Pugh class and discovered that patients with Child– Pugh class A classification exhibited a significant reduction in major bleeding risk in DOAC users (aHR, 0.48; 95% CI, 0.33–0.70); however, this reduction was nonsignificant for patients with class B or C classification (aHR, 0.77; 95% CI, 0.54−1.08) (**Figure 3A and 3B**).

**Figure 3.**
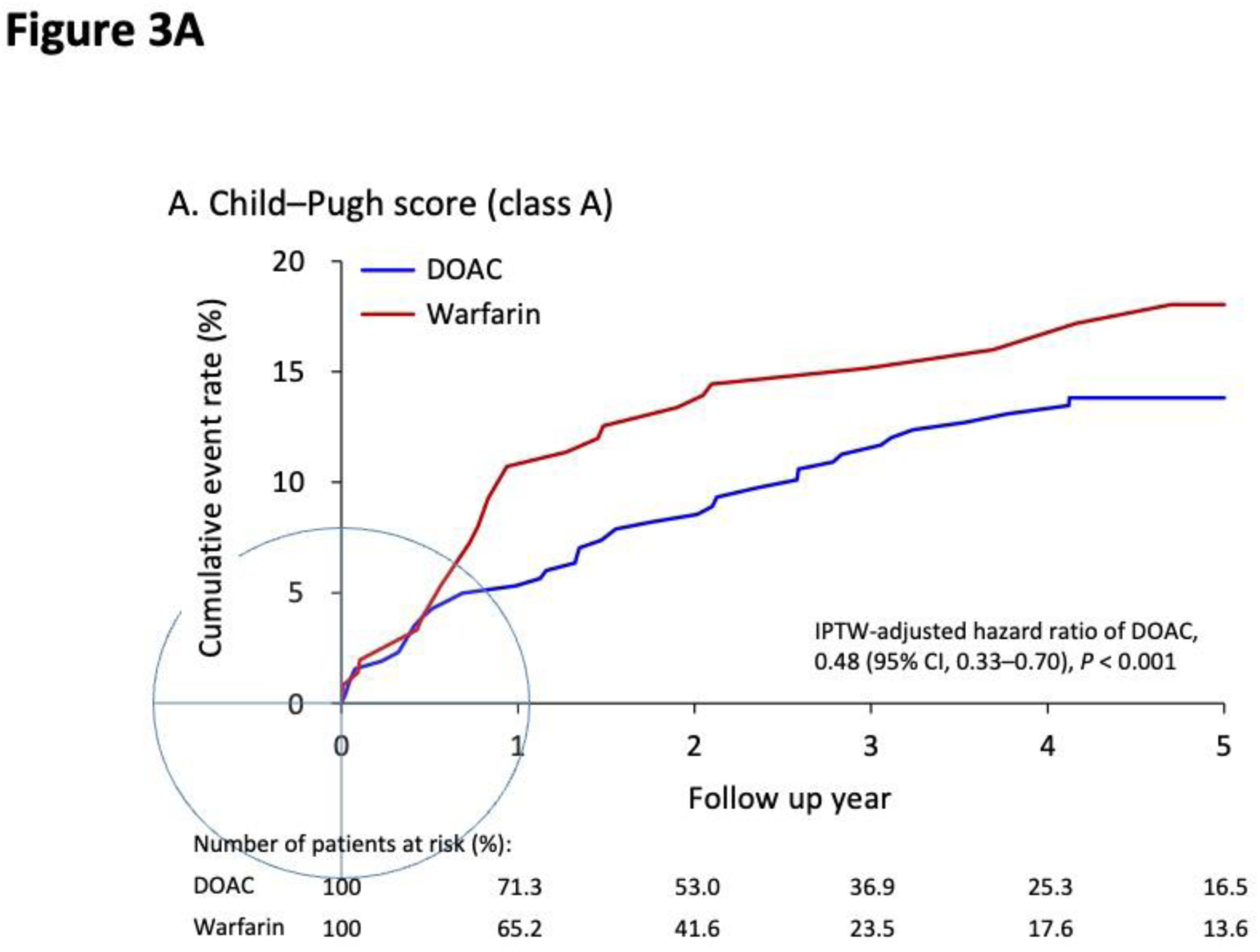
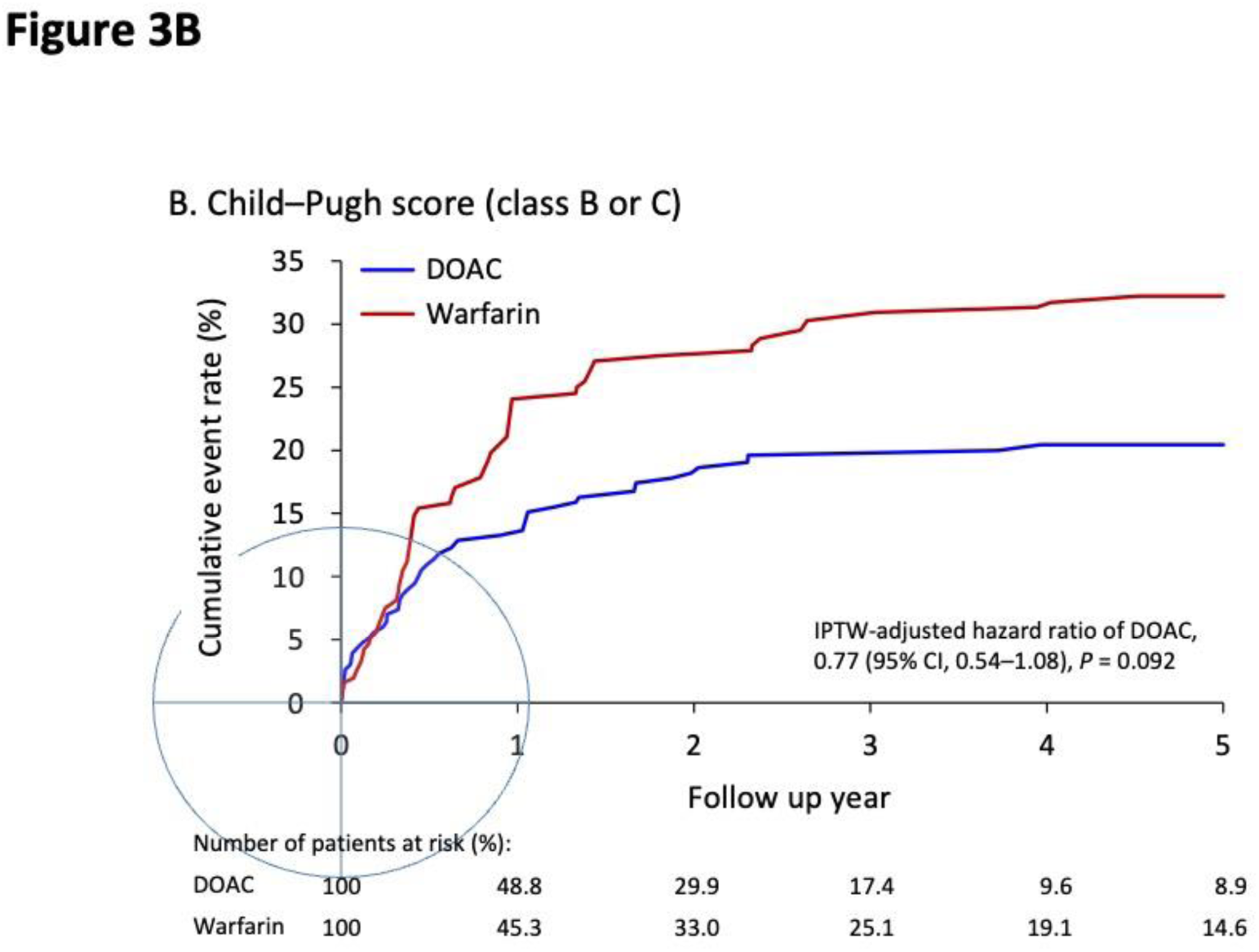
Cumulative event rates for primary safety outcome (major bleeding) during 5-year follow-up period for patients with atrial fibrillation and liver cirrhosis who were administered DOACs or warfarin in IPTW-adjusted cohort; results are classified under Child–Pugh class A (A) and Child–Pugh class B or C (B). Abbreviations: DOAC, direct oral anticoagulant; IPTW, inverse probability of treatment weighting.

## Discussion

In the present cohort study, we evaluated the efficacy and safety of DOACs versus warfarin among 725 patients with non-valvular AF and LC of varying severity. Our findings indicated that DOACs and warfarin were similarly effective in preventing thromboembolic events, namely IS, TIA, and SE. Compared with warfarin users, DOAC users exhibited similar ICH risk and a non-significantly lower risk of GI bleeding. DOAC use was associated with a lower risk of major bleeding, especially in patients with Child–Pugh class A LC. DOAC use was associated with lower all-cause mortality. Regarding liver decompensation outcomes, no significant differences were observed between the DOAC and warfarin groups. Overall, DOAC and warfarin users exhibited similar efficacy and safety with respect to ICH and GI bleeding. However, relative to warfarin, DOACs exhibited superior safety in reducing major bleeding events; this finding provides essential guidance when deciding whether to administer anticoagulation therapy for patients with AF and LC.

For patients with LC, DOACs and warfarin have been reported to lead to similar outcomes with respect to stroke and SE (17–19). In our study, patients with LC who used DOACs exhibited similar levels of IS and SE risk relative to those who used warfarin. The pathophysiological mechanism underlying the association between LC and stroke is complex and multifactorial. Clinical evidence has debunked the misconception that natural anticoagulation occurs in patients with LC (25). Notably, the risk of ischemic stroke has been reported to be higher in patients with AF and LC than in patients with AF alone (26). Chronic liver disease, particularly LC, increases thrombosis risk because of the impaired synthesis of endogenous anticoagulants such as protein C, protein S, and antithrombin (25). This risk is intensified because of the combined effects of several factors in patients with LC, namely the release of factor VIII and von Willebrand factor stimulated by lipopolysaccharides from bacterial translocation; the reduction in ADAMTS13 levels, which modulates von Willebrand factor activity; and hypofibrinolysis due to low plasminogen levels (27, 28). Furthermore, in patients with LC, interleukin-6, which is frequently elevated in response to endotoxemia (29, 30), increases tissue factor expression on activated monocytes and endothelial cells (31). This cascade amplifies systemic inflammation, impairs the coagulation pathway, and potentially increases the risk of stroke in patients with LC (32, 33). In our study cohort, these LC-related factors could have precipitated stroke and reduced the protective effect of DOACs, possibly explaining the absence of a significant effect on stroke outcomes.

Compared with warfarin use, DOAC use was reported to be associated with a lower risk of ICH in several key randomized controlled trials (8–11). In the present study, a similar level ICH risk was also observed among DOAC users. A proposed hypothesis is that DOACs, by not interfering with the formation of tissue factor and factor VIIa complexes, may support cerebral hemostatic mechanisms (34). Additionally, the interactions of DOACs with the P-glycoprotein transport system and their potential ability to limit endothelial cell damage within the blood–brain barrier (mediated by the thrombin/Protease-Activated Receptor-1 pathway) may contribute to their superior safety profile in preventing ICH (34–36). Another plausible hypothesis is that anticoagulants disrupt the coagulation pathway, which promotes the accelerated extravasation of red blood cells, the accumulation of cerebral microhemorrhages (CMBs), and, eventually, leads to the development of symptomatic ICH (37). This hypothesis is supported by a study that reported a lower incidence of new CMB among DOAC users than among warfarin users in a population treated for AF (38). However, findings have been inconsistent regarding the association of ICH risk with DOAC use in the context of liver disease (17, 18). Increased liver fibrosis may be independently associated with an increased burden of CMB, but the mechanism underlying this association remains unclear (39). Furthermore, patients with LC exhibit an increased risk of hemorrhagic stroke (40). Although DOACs can marginally reduce the risk of ICH in patients without LC, the complex nature of ICH development may diminish this protective effect in patients with LC. This supports our finding that DOAC and warfarin users exhibit similar levels of ICH risk.

The present study revealed that relative to warfarin users, DOAC users tended to exhibit a reduced risk of GI bleeding and had a decreased risk of major bleeding. The superior safety profiles of DOACs relative to warfarin may be attributed to their more predictable pharmacokinetics, lower dependence on liver clearance, and fewer interactions with various drugs and foods. Furthermore, bacterial overgrowth in the small intestine, which is commonly observed in patients with LC, particularly those with advanced liver disease and portal hypertension, may alter gut microbiota (41, 42). This change can lead to a net decrease in vitamin K availability, which may amplify the anticoagulant effect of warfarin, thereby increasing the risks of over-anticoagulation and bleeding (43). The aforementioned factors may have collectively contributed to the observed reduction in the risks of major and GI bleeding. Compared with the cohorts of other studies, our cohort exhibited several distinct differences. First, our cohort had a lower HAS-BLED score relative to that of a nationwide U.S. study that included not only patients with LC but also those with less severe liver diseases (19). Despite the lower HAS-BLED score, our study revealed higher risks of both major and GI bleeding. This finding indicates that the HAS-BLED score should be further validated to more accurately predict the bleeding risk associated with anticoagulant use in patients with LC. Second, Lawal et al. categorized patients with LC into those with compensated or decompensated LC, and they did not identify any differences in clinical outcomes (including major and GI bleeding) between DOAC and warfarin users (19). Conversely, our analysis highlighted the safety benefits of DOAC, particularly with respect to major bleeding, for patients with Child–Pugh class A LC. Additionally, DOACs demonstrated a trend towards a reduction in GI bleeding risk in our cohort, although this trend did not reach statistical significance.

In our study, the risk of all-cause mortality was lower among DOAC users than among warfarin users, and this trend was most pronounced for liver-related deaths (Supplemental Figure S1). While a study found that the mortality benefit of DOAC may be related to the reduced risk in ICH (44), our study observed a comparable risk of ICH with DOAC use. This suggests that the advantage of DOAC in reducing all-cause mortality may be partly due to a decrease in liver-associated complications, though the underlying biological mechanisms for this effect remain to be elucidated. A study highlighted the mortality-reducing effects of vitamin K1 in patients with LC (45), providing a backdrop for exploring the interplay between anticoagulants and vitamin K. Additionally, a meta-analysis revealed that patients experiencing ICH while on DOACs had smaller hematoma sizes and lower mortality rates at discharge compared to those on vitamin K antagonists (46). This indicates that DOACs are associated with less severe outcomes in the event of ICH, potentially contributing to their overall mortality benefit. Furthermore, nearly half of our study participants were diagnosed with diabetes mellitus. The benefits of vitamin K in enhancing insulin sensitivity and mitigating inflammation raises questions about whether warfarin’s inhibition of vitamin K may exacerbate glucose control and inflammatory responses (47). A supporting study reported lower mortality rates among DOAC users than among warfarin users in patients with AF and diabetes mellitus (48). Nevertheless, the aforementioned hypotheses represent preliminary interpretations, and more rigorous research is required to confirm them.

### Limitations

Our study has several limitations: 1. **Patient Cohort**: Our cohort consists mostly of patients with milder forms of LC, which limits our findings’ applicability to advanced LC (Child–Pugh classes B and C). However, the relevance of stroke prevention for patients with Child–Pugh class B and C LC, who often have a limited life expectancy, may be reduced. Thus, the long-term effects of DOACs in these classes may be less crucial. 2. **Differential Effects of DOACs**: Although each DOAC has a distinct pharmacological profile, our study did not separately examine the safety and efficacy of each DOAC in this cohort. Notably, a study reported a higher bleeding risk in rivaroxaban users than in apixaban users (49). Because of the limited number of participants in each DOAC class, our study did not have adequate statistical power for the further analysis of each DOAC. 3. **Methodological Limitation**: We employed IPTW to mitigate biases when comparing DOACs and warfarin. However, retrospective studies inherently harbor confounders, which hinder the confirmation of causality between DOAC use and outcomes, and only randomized controlled trials or prospective studies can yield conclusive evidence. 4. **Generalizability**: The present study’s geographical context and predominant LC etiology restrict its general applicability. Specifically, it was conducted within the context of an East Asian health-care system where viral hepatitis and alcohol are the primary causes of LC. Therefore, our results may not be generalized to patients with liver diseases resulting from nonalcoholic steatohepatitis, although literature findings suggest that the pharmacokinetics of apixaban are unaffected by nonalcoholic steatohepatitis (50).

### Conclusion

In the present study, DOACs and warfarin demonstrated comparable effectiveness in preventing thromboembolic events, namely IS, TIA, and SE. However, compared to warfarin, DOAC use resulted in lower rates of major bleeding, particularly in patients with Child–Pugh class A LC, while demonstrating comparable effects on the risks of ICH and GI bleeding. DOACs were associated with reduced all-cause mortality. Our research provides a real-world perspective that can help to guide clinicians in anticoagulation decision-making for patients with AF and LC.

## Data Availability

Upon Request

## Acknowledgments

The authors thank the statistical assistance and wish to acknowledge the support of the Maintenance Project of the Center of data science and Biostatistics (Grant CGRPG2F0011, CLRPG2C0021, CLRPG2C0022, CLRPG2C0023, CLRPG2C0024, CLRPG2G0081, CLRPG2G0082, CLRPG2G0083, CLRPG2L0021, CLRPG2L0022, and CMRPG2N0041) at Keelung Chang Gung Memorial Hospital for study design and monitor, data analysis and interpretation.

